# Adverse Childhood Events, Mood and Anxiety Disorders, and Substance Dependence: Gene X Environment Effects and Moderated Mediation

**DOI:** 10.1101/2023.10.24.23297419

**Authors:** Henry R. Kranzler, Christal N. Davis, Richard Feinn, Zeal Jinwala, Yousef Khan, Ariadni Oikonomou, Damaris Silva-Lopez, Isabel Burton, Morgan Dixon, Jackson Milone, Sarah Ramirez, Naomi Shifman, Daniel Levey, Joel Gelernter, Emily E. Hartwell, Rachel L. Kember

## Abstract

**Background:** Adverse childhood events (ACEs) contribute to the development of mood and anxiety disorders and substance dependence. However, the extent to which these effects are direct or indirect and whether genetic risk moderates them is unclear. **Methods:** We examined associations among ACEs, mood/anxiety disorders, and substance dependence in 12,668 individuals (44.9% female, 42.5% African American/Black, 42.1% European American/White). We generated latent variables for each phenotype and modeled direct and indirect effects of ACEs on substance dependence, mediated by mood/anxiety disorders (forward or “self-medication” model) and of ACEs on mood/anxiety disorders, mediated by substance dependence (reverse or “substance-induced” model). In a sub-sample, we also generated polygenic scores for substance dependence and mood/anxiety disorder factors, which we tested as moderators in the mediation models. **Results:** Although there were significant indirect effects in both directions, mediation by mood/anxiety disorders (forward model) was greater than by substance dependence (reverse model). Greater genetic risk for substance dependence was associated with a weaker direct effect of ACEs on substance dependence in both the African- and European-ancestry groups (i.e., gene-environment interaction) and a weaker indirect effect in European-ancestry individuals (i.e., moderated mediation). **Conclusion:** We found greater evidence that substance dependence results from self-medication of mood/anxiety disorders than that mood/anxiety disorders are substance induced. Among individuals at higher genetic risk for substance dependence who are more likely to develop a dependence diagnosis, ACEs exert less of an effect in promoting that outcome. Following exposure to ACEs, multiple pathways lead to mood/anxiety disorders and substance dependence. Specification of these pathways could inform individually targeted prevention and treatment approaches.

## INTRODUCTION

Adverse childhood events (ACEs)—such as childhood sexual abuse, physical abuse, and exposure to violence or parental alcohol and/or drug abuse—have consistently been shown to contribute to the development of psychiatric disorders, including major depressive disorder (MDD), post-traumatic stress disorder (PTSD), and substance use disorders (SUDs)^1–12^. There is a strong linear relationship between the number of ACEs individuals experience and their risk for adverse health outcomes^13^. In a latent class analysis (LCA) of the effects of exposure to 13 different ACEs (e.g., maltreatment, household dysfunction, and community violence), after controlling for sociodemographic characteristics and common risk factors for substance use, young adults in the high/multiple ACEs class reported higher levels of alcohol-related problems, current tobacco use, and psychological symptoms than those in the low ACEs class^10^. Similarly, in an LCA of 11,386 US older adults that yielded 4 classes of ACEs—high adversity (6%), low adversity (69%), child abuse (16%), and parental substance abuse (8%)—rates of SUDs were lower in the low-adversity group than the other three groups after controlling for demographic measures. Additionally, the high-adversity and child abuse groups were more likely to have experienced major depression than the low-adversity group.^12^

Greater insight into the nature of the relationship between ACEs and mood/anxiety and SUDs can be gained from mediation models, which can differentiate direct effects of ACEs on these disorders from indirect (i.e., mediated) ones and show the directionality of the causal effects, which can both inform theory and have clinical utility. Douglas et al.^14^ reported that a summary measure of mood and anxiety disorders (M/ADs) that preceded a substance dependence (SD) diagnosis partially mediated the effect of ACEs on SD risk.

Here, we sought to replicate those findings in a larger sample and to compare two mediation models that differed in the hypothesized direction of effect by using information on ages of onset of the disorders to establish temporality. The forward model, consistent with the self-medication hypothesis^15–17^, posits that M/ADs mediate the relationship between ACEs and SD diagnoses. In the reverse model, consistent with M/ADs being substance induced^18^, SD mediates the relationship between ACEs and M/ADs. In view of the bidirectional relationship between M/ADs and SD described in the literature ^19–31^, we examined effects in both directions.

Whereas not everyone who experiences an ACE goes on to develop an M/AD or SD, genetic liability could help to explain variation in risk for these disorders^32^. Incorporating genetic information into mediation analyses can indicate whether causal pathways in the development of SUDs and M/ADs differ for individuals at varying levels of genetic risk (i.e., whether there exists a gene x environment (GxE) interaction). Specifying the pathway from early childhood experiences to later psychiatric and substance-related outcomes is critical for developing effective, individually targeted prevention and treatment approaches. Polygenic risk scores (PRS), calculated by summing the effects of genetic variants across the genome, can reflect an individual’s aggregate genetic liability for a particular trait or disorder. The effects of PRS, if large enough, could be useful clinically^33–35^. We therefore also examined PRS for M/ADs and SD as moderators of the direct and indirect effects of ACEs in both models.

We tested several hypotheses. First, we expected to replicate the findings of Douglas et al.^14^ by showing that ACEs have both direct and indirect effects on SD risk, with the latter mediated by M/ADs. Second, we hypothesized that this mediating effect of M/ADs in the forward, self-medication model would be larger than the mediating effect of an SD diagnosis on the relationship between ACEs and the occurrence of an M/AD in the reverse, substance-induced model. Third, we hypothesized that the SD PRS would positively moderate the direct effect of ACEs on SD (i.e., that a higher SD PRS would be associated with a greater impact of ACEs on SD). Fourth, we hypothesized that the M/AD PRS would positively moderate the direct effect of ACEs on M/AD (i.e., that a higher M/AD PRS would be associated with a greater impact of ACEs on M/ADs). Finally, we hypothesized that the SD and M/AD PRS would moderate the mediating effects of M/ADs and SD, respectively, with no directional hypotheses for these effects.

## METHODS

### Overview

We first examined the subgroup of participants who had onset of an M/AD prior to the onset of an SD diagnosis (forward model consistent with a self-medication hypothesis) and then repeated the analysis among individuals whose first SD diagnosis occurred prior to the onset of an M/AD (reverse model consistent with substance-induced mood changes). The mediation analyses combined all individuals irrespective of self-identified race. We used summary statistics from large genome-wide association studies (GWAS) of M/ADs,^36–44^ and SUDs^45–49^ to derive latent genetic factors for the two traits using genomic structural equation modeling (gSEM). Using summary statistics from GWAS on the latent genetic factors, we calculated PRS in the Yale-Penn sample. We then examined the moderating effect of PRS for M/ADs on the relationship between ACEs and M/ADs and of PRS for SD on the relationship between ACEs and SD, separately by ancestral group.

### Participants

We included 12,668 individuals from the Yale-Penn sample, a family-based and case-control sample that was recruited at Yale University, UConn Health, the University of Pennsylvania, the Medical University of South Carolina, and McLean Hospital for studies of the genetics of SD^50^. The institutional review board at each site approved the study protocol. All participants received a complete description of the study, gave written informed consent, and were paid to complete the assessments and provide a blood or saliva sample for genotyping.

### Assessment Procedures

The family-based sample was ascertained through two or more siblings affected with a lifetime diagnosis of cocaine and/or opioid dependence. The case-control sample comprises unrelated cases with a lifetime diagnosis of alcohol, cocaine, tobacco, cannabis, or opioid dependence, and controls who were included based on the absence of any of these diagnoses. Additional family members of probands from family-based studies and of cases from case-control studies were also recruited irrespective of their SD status. DSM-IV^51^ SD diagnoses were obtained with the Semi-Structured Assessment for Drug Dependence and Alcoholism (SSADDA), which elicits information on age of onset of a variety of psychiatric and SD diagnoses. The reliability of the SSADDA for both psychiatric and SD diagnoses and individual criteria has been reported previously^52,53^. Whereas SD diagnoses in the Yale-Penn sample were based on DSM-IV, diagnoses in the GWAS discovery samples used to generate PRS were largely based on SUD diagnoses in the International Classification of Diseases.

### Measures

<u>Adverse Childhood Events Latent Variable.</u> Ten variables reflected participants’ experiences in before age 13. The test-retest and inter-rater reliability estimates for these items ranged from 0.62 to 0.99 and 0.41 to 0.82, respectively. All variables were dichotomized to ensure the same coding and equal weight among them. Two variables reflected the perceived instability of the participants’ home life: multiple main caregivers (3 or more) and multiple family relocations (2 or more). Three variables reflected childhood traumatic experiences: violent crime, sexual abuse, and physical abuse. Violent crime was defined as witnessing or experiencing a violent crime, like a shooting or a rape. Sexual abuse was assessed by asking whether the respondent was ever sexually abused. Physical abuse was defined as being beaten by an adult so badly that medical care was needed or marks on the body remained for more than 30 days. The remaining variables were household substance use, regular household smoking, and three variables thought to be potentially protective against ACEs that were reverse coded so that lower levels were less protective: frequency of religious participation (never vs. ever), quality of relationship with the main caregiver (poor vs. moderate or greater), and frequency of contact with other relatives (less than monthly vs. more frequent contact). All 10 variables loaded significantly onto a single ACEs latent variable, with item loadings ranging from 0.16 (for no religious involvement) to 0.71 for physical abuse (see Supplementary Figure 1).

<u>Substance Dependence Latent Variable.</u> The SD latent variable comprised DSM-IV SD diagnoses for alcohol, cocaine, opioids, tobacco, and cannabis. These five diagnoses loaded well onto a single factor, with all item loadings ≥0.69 (see Supplementary Figure 2).

<u>Mood and Anxiety Disorders Latent Variable.</u> We included eight psychiatric disorders (major depressive disorder [MDD], bipolar disorder, posttraumatic stress disorder [PTSD], generalized anxiety disorder [GAD], obsessive-compulsive disorder [OCD], social phobia, agoraphobia, and panic disorder) as indicators for a single M/AD latent variable. We included PTSD among the M/ADs because diagnoses in the target sample were made using DSM-IV, in which PTSD is categorized as an anxiety disorder. All item loadings were significant, ranging from 0.11 for MDD to 1.00 for PTSD (see Supplementary Figure 3).

<u>Demographics.</u> Seven demographic characteristics served as control variables in the mediation models: sex, age, self-identified race/ethnicity [African American/Black, European American/White, Hispanic/Latino, or Other (includes Native/American, Asian, Pacific Islander, and Other)], marital status (never married vs. ever married or cohabiting), years of education, employment status, and annual household gross income (≤$10,000, $10–29,999, $30-74,999, ≥$75,000). Self-identified race/ethnicity was omitted as a control variable in the moderation analyses, which were run separately for the African- and European-ancestry groups.

<u>Polygenic Risk Scores.</u> To create PRS that captured general genetic liability for M/ADs and SUDs, we used summary statistics from large genome-wide association studies (GWAS) of M/ADs (Supplementary Tables 1 and 2) and SUDs (Supplementary Tables 3 and 4) and genomic structural equation modeling (gSEM^54^) to derive latent genetic factors for African- and European-ancestry separately (Supplementary Tables 5-10 and Supplementary Figures 4-7 provide additional details). Prior to applying gSEM, multi-trait analysis of GWAS (MTAG^55^) was used to jointly analyze summary statistics from three GWAS of anxiety-related traits in European-ancestry samples^36–38^ to boost power to detect genetic associations related to a broad spectrum of anxiety disorders. Using GWAS of the latent genetic factors, we generated summary statistics and then calculated ancestry-specific M/AD and SUD PRS in the Yale-Penn sample (African-ancestry n = 2,284, European-ancestry n = 2,871; details on genetic QC and ancestry assignment have been reported previously^50^.

### Analyses

We compared demographic characteristics across SD and M/AD groups using chi-square for categorical variables and t-test for continuous variables. We also examined whether the odds of experiencing each of the ACEs differed for participants with and without SD and M/AD diagnoses using Mplus version 8.10^56^. First, latent factors were created for ACEs, SD, and M/AD. Whereas all indicators were binary variables, we used the WLSMV estimator. The factor loadings of all indicators in the correlated factors CFA were significant, and the overall model demonstrated good fit (CFI=0.95, RMSEA=0.037). Next, the factor loadings were fixed for the two mediation models (see Figure 1), one in which the M/AD factor was the mediator and the SD factor the outcome (i.e., the forward or self-medication model) and a second model in which the mediator was the SD factor and the M/AD factor the outcome (i.e., the reverse or substance-induced model). In both models, the ACEs factor served as the independent variable. Age of onset for first SD diagnosis and first M/AD was used to select a subset of the sample for each mediation model to maintain the temporal order of mediation.

**Figure 1:**
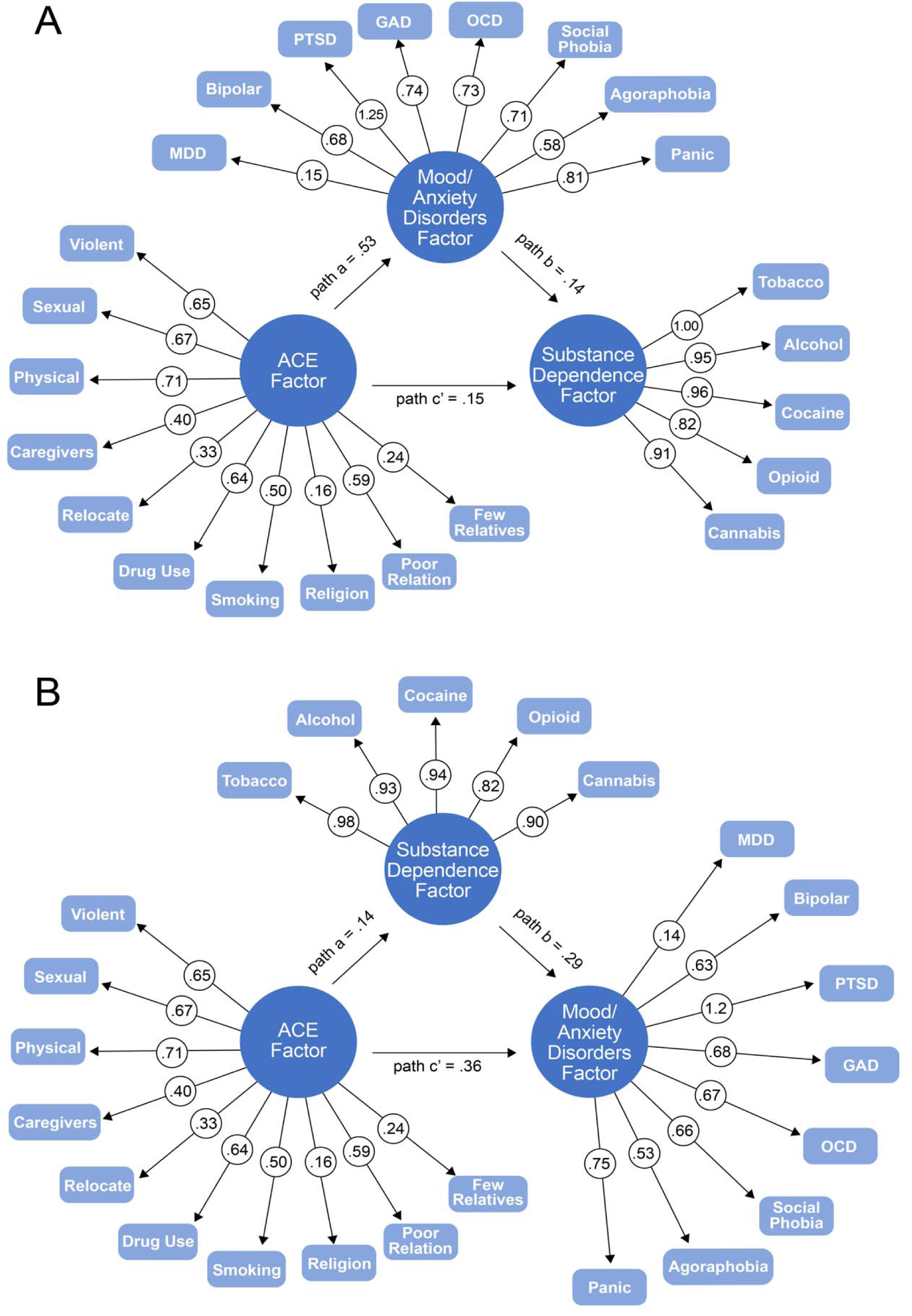
Path diagrams of the mediation models. Panel A shows the forward mediation model and Panel B the reverse mediation model. All paths are significant (p<0.05).

Excluding participants from the forward model if the onset of their first SD occurred before the onset of their first M/AD left a final n = 4,128. Similarly, excluding participants from the reverse model if the onset of their first M/AD occurred before the age of onset for SD left a final n = 2,957. Participants who had the same age of onset for SD and an M/AD (n = 395) were included in both models. As a sensitivity analysis, we repeated the mediation analyses after excluding these individuals. Whereas the results were substantively unchanged, we present the findings that include these individuals.

The mediator and outcome in both models were regressed on the demographic covariates. To account for the fact that 2% of participants belonged to the same family, we used the COMPLEX analysis option for nested data. The model INDIRECT command was used to estimate the indirect effects and the bootstrap procedure with 1,000 bootstrap samples served to generate 95% confidence intervals.

We then conducted moderated mediation analyses that included the SUD and M/AD PRS as moderators. In the forward or self-medication models (see Figure 2 Panel A), we included the M/AD PRS as a moderator of the indirect path from ACEs to M/ADs (path a), the SUD PRS as a moderator of the indirect path from M/ADs to SD (path b), and SUD PRS as a moderator of the direct path from ACEs to SD (path c’). In the reverse or substance-induced models (see Figure 2 Panel B), we included the SUD PRS as a moderator of the indirect path from ACEs to SD (path a), the M/AD PRS as a moderator of the indirect path from SD to M/ADs (path b), and the M/AD PRS as a moderator of the direct path from ACEs to M/ADs (path c’). The moderated mediation analyses were limited to the subset of participants with genetic data (African ancestry, n = 2,284; European ancestry, n = 2,871) and were run separately by ancestral group.

**Figure 2:**
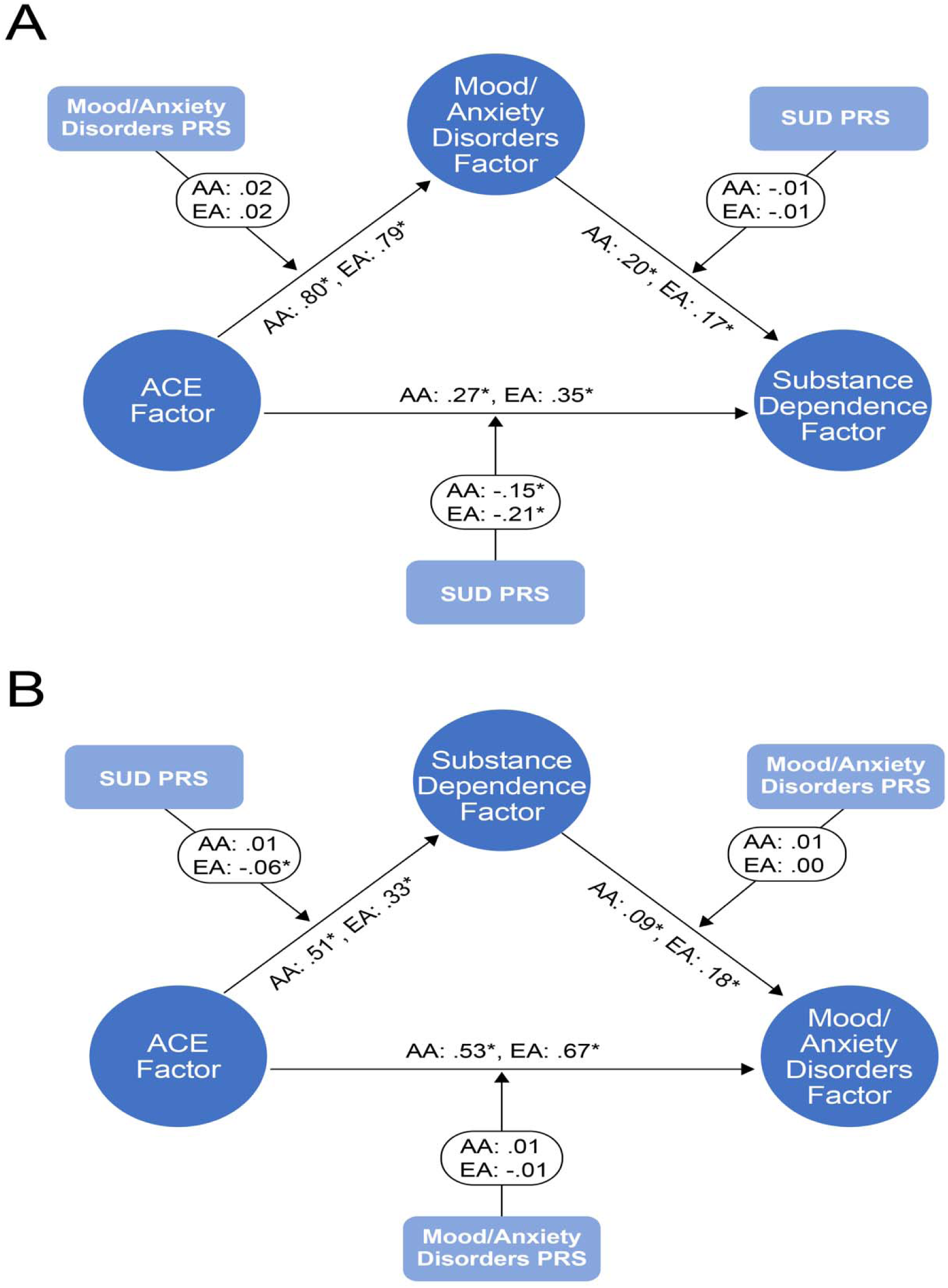
Path diagrams of the moderated mediation model results. *Significant paths. Panel A presents the forward moderated mediation model and Panel B the reverse moderated mediation model. AA = African ancestry, EA = European ancestry, SUD PRS = substance use disorders polygenic risk score.

## RESULTS

As shown in Table 1, the Yale-Penn sample comprised 12,668 individuals, 55.1% of whom were male, with an average age of 40.7 years (SD = 12.1) and nearly equal percentages of self-identified African American/Black (42.5%) and European American/White (42.1%), and smaller percentages of Hispanic/Latino (7.6%) and other racial/ethnic groups (7.7%). Only 17.8% of the sample was married or cohabiting. Participants averaged 12.8 (SD = 2.4) years of education, and 57.1% were currently employed. A plurality of individuals (43%) reported an annual gross household income of less than $10,000.

**Table 1.** Demographic characteristics of participants with and without a substance dependence diagnosis.

|  | Overall<br>N=12,668 | Substance<br>Dependence<br>N=9,695 | No Substance<br>Dependence<br>N=2,973 | p-value | Effect Size* |
| --- | --- | --- | --- | --- | --- |
| **Sex** |  |  |  | <0.001 | 0.578 |
| Male | 55.1% | 61.7% | 33.8% |  |  |
| **Age** |  |  |  | <0.001 | 0.076 |
| Mean ± std. dev. | 40.7 ± 12.1 | 40.5 ± 11.2 | 41.4 ± 14.6 | | |
| **Race/Ethnicity** |  |  |  | <0.001 | 0.127 |
| African American | 42.5% | 43.6% | 39.2% |  |  |
| European American | 42.1 | 40.5 | 47.5 |  |  |
| Hispanic/Latino | 7.6 | 8.1 | 5.9 |  |  |
| Other | 7.7 | 7.8 | 7.5 |  |  |
| **Marital Status** |  |  |  | <0.001 | 0.626 |
| Married | 17.8% | 12.4% | 35.6% |  |  |
| **Education (Years)** |  |  |  | <0.001 | 1.211 |
| Mean ± std. dev. | 12.8 ± 2.4 | 12.2 ± 2.2 | 14.7 ± 2.0 | | |
| **Employment** |  |  |  | <0.001 | 0.637 |
| Employed | 57.1% | 48.5% | 78.8% |  |  |
| **Income** |  |  |  | <0.001 | 1.043 |
| <\$10,000 | 43.0% | 51.3% | 15.6% | | |
| \$10,000-\$29,999 | 28.2 | 29.4 | 24.2 | | |
| \$30,000-\$74,999 | 20.4 | 14.9 | 39.5 | | |
| \$75,000+ | 8.4 | 4.4 | 21.6 | | |
| **Substance Dependence** |  |  |  |  |  |
| Tobacco |  | 81.0% |  |  |  |
| Alcohol |  | 88.5% |  |  |  |
| Cocaine |  | 73.2% |  |  |  |
| Opioid |  | 41.9% |  |  |  |
| Marijuana |  | 60.1% |  |  |  |
| Mean # of diagnoses ± std. dev. | | 3.2 ± 1.3 | | | |

*Note:* \* Standardized difference with eta-squared conversion applied to race. std. dev. = standard deviation.

There were 9,695 cases (76.5%) with one or more lifetime alcohol, cocaine, opioid, tobacco, or cannabis dependence diagnoses, and 2,973 SD controls (23.5%) (see Table 2). Nearly one-third of individuals (4,205 or 33.2%) had one or more M/ADs and the remaining 8,463 participants (66.8%) had no M/AD. Nearly half (6,032 or 47.6%) of participants had only an SD diagnosis, 542 (4.3%) had only an M/AD diagnosis, 3,663 (28.9%) had both SD and M/AD diagnoses, and 2,431 (19.2%) had neither diagnosis.

**Table 2.**
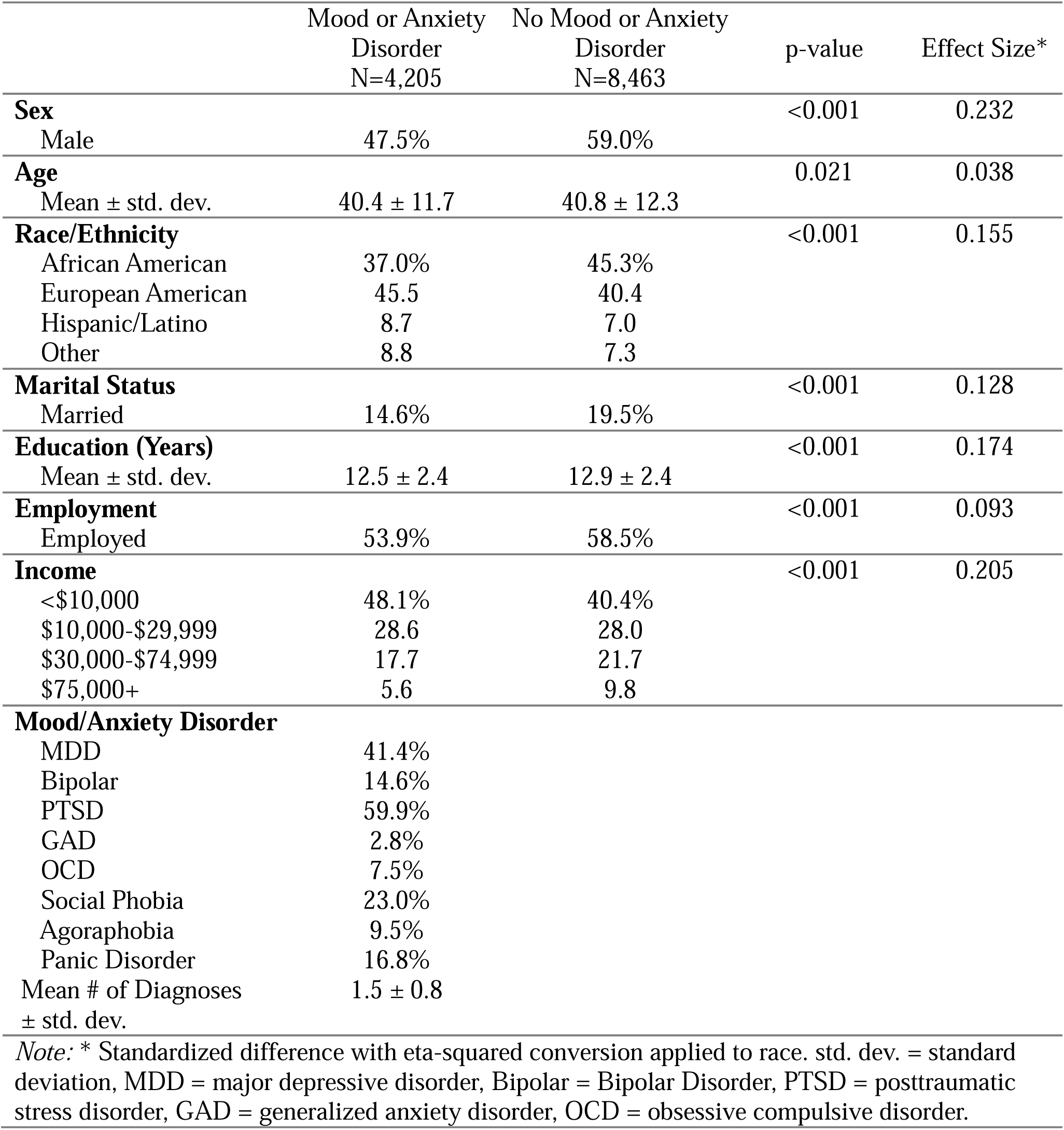
Demographic characteristics of participants with and without a mood or anxiety disorder.

The most common SD diagnosis was alcohol dependence (89%) and the least common was opioid dependence (42%). The average number of SD diagnoses was 3.2 (SD = 1.3), ranging from one (12%) to five (17%). Participants with an SD diagnosis were younger and more likely to be male and African American, have lower income and a lower education level, and less likely to be employed or married (see Table 1). Participants with SD were more likely to experience each of the ACEs and to lack protective factors (see Table 3), with the highest odds being for physical abuse (OR = 4.16), witnessing violent crime (OR = 3.91), and household substance use (OR = 3.56).

**Table 3.** Prevalence rates and odds of adverse childhood events across substance dependence and mood and anxiety disorder groups.

|  | Substance<br>Dependence | No<br>Substance<br>Dependence | OR | Mood/<br>Anxiety<br>Disorder | No<br>Mood/Anxiety<br>Disorder | OR |
| --- | --- | --- | --- | --- | --- | --- |
| *Adverse Childhood Events* |  |  |  |  |  |  |
| Violent Crime | 21.0% | 6.4% | 3.91 | 26.1% | 13.2% | 2.32 |
| Sexual Abuse | 17.0% | 7.3% | 2.61 | 26.1% | 9.0% | 3.57 |
| Physical Abuse | 10.4% | 2.7% | 4.16 | 15.9% | 5.0% | 3.59 |
| 3+ Main Caregivers | 7.7% | 4.6% | 1.73 | 10.1% | 5.3% | 2.00 |
| 2+ Relocations | 55.5% | 43.4% | 1.63 | 58.1% | 49.9% | 1.39 |
| Household Substance<br>Use | 59.3% | 29.0% | 3.56 | 62.5% | 46.9% | 1.89 |
| Household Smoking | 75.0% | 53.1% | 2.65 | 76.6% | 66.4% | 1.66 |
| No Religious<br>Participation* | 11.0% | 8.8% | 1.28 | 11.4% | 10.0% | 1.16<sup>a</sup> |
| Poor Caregiver<br>Relationship* | 6.7% | 3.1% | 2.29 | 11.1% | 3.3% | 3.72 |
| Infrequent Contact<br>with Relatives* | 6.4% | 3.5% | 1.89 | 6.6% | 5.3% | 1.28<sup>b</sup> |

*Note:* \*Refers to lack of protective factors against the harms of adverse childhood events; SD = substance dependence, OR = odds ratio. All p-values are <.001 unless otherwise indicated. <sup>a</sup>p-value = .014, <sup>b</sup>p-value = .002.

The most common M/AD diagnosis was PTSD (59.9%), with generalized anxiety disorder being least prevalent (3%). The average number of M/ADs was 1.5 (SD=0.8), with most participants (68%) having a single diagnosis. Individuals with an M/AD diagnosis were younger and more likely to be female, European American, have a lower income, and less likely to be married or working. Individuals with an M/AD diagnosis were more likely to experience ACEs and to lack protective factors (see Table 3), with the largest effects being that of a poor relationship with the main caregiver (OR = 3.72), experiencing physical abuse (OR = 3.59), and experiencing sexual abuse (OR = 3.57). Non-participation in religious activities had the weakest association with both outcomes (SD: OR = 1.28, M/ADs: OR = 1.16).

### Mediation Models

All paths in the forward mediation model were significant. There was a direct effect of ACEs on SDs (β = 0.15, SE = 0.01, *p* < 0.001), with individuals who experienced more ACEs being more likely to develop an SD. There was also an indirect effect operating through M/ADs (β = 0.08, SE = 0.01, *p* < 0.001; see Table 4 and Figure 1, Panel A), which accounted for 34% (0.075/0.221) of the total effect of ACEs on SD.

**Table 4.** Results of mediation models.

| | $\beta$ | SE | p-value | 95% CI |
| --- | --- | --- | --- | --- |
| ***Forward Model: Adverse Childhood Events (ACEs)→Mood and Anxiety Disorders (M/ADs)→Substance Dependence (SD)<sup>1</sup>*** |  |  |  |  |
| ACEs→M/ADs | 0.527 | 0.009 | <0.001 | 0.507 – 0.543 |
| M/ADs→SD | 0.143 | 0.008 | <0.001 | 0.129 – 0.160 |
| ACEs→SD | 0.145 | 0.009 | <0.001 | 0.128 – 0.161 |
| Indirect Effect |  |  |  |  |
| ACEs→M/ADs→SD | 0.075 | 0.005 | <0.001 | 0.067 – 0.085 |
| Direct Effect |  |  |  |  |
| ACEs→SD | 0.145 | 0.009 | <0.001 | 0.128 – 0.161 |
| Total Effect |  |  |  |  |
| Indirect + Direct | 0.221 | 0.008 | <0.001 | 0.205 – 0.237 |
| ***Reverse Model: Adverse Childhood Events (ACEs)→Substance Dependence (SD)→Mood and Anxiety Disorders (M/ADs)<sup>2</sup>*** |  |  |  |  |
| ACEs→SD | 0.144 | 0.011 | <0.001 | 0.122 – 0.166 |
| SD→M/ADs | 0.285 | 0.031 | <0.001 | 0.232 – 0.352 |
| ACEs→M/ADs | 0.356 | 0.033 | <0.001 | 0.282 – 0.381 |
| Indirect Effect |  |  |  |  |
| ACEs→SD→M/ADs | 0.041 | 0.005 | <0.001 | 0.032 – 0.052 |
| Direct Effect |  |  |  |  |
| ACEs→M/ADs | 0.356 | 0.033 | <0.001 | 0.282 – 0.381 |
| Total Effect |  |  |  |  |
| Indirect + Direct | 0.397 | 0.031 | <0.001 | 0.326 – 0.421 |
| *Note:* Models included sex, age, race, marital status, years of education, employment status, and income as covariates. SE = standard error, CI = confidence interval. |  |  |  |  |
| <sup>1</sup>Model was restricted to participants with an age of onset of first substance dependence diagnosis at or after their age of onset for a first mood/anxiety disorder diagnosis. |  |  |  |  |
| <sup>2</sup>Model was restricted to participants with an age of onset for a first mood/anxiety disorder at or after their age of onset for a first substance dependence diagnosis. |  |  |  |  |

All paths in the reverse model were also significant (see Table 4 and Figure 1, Panel B). There was a significant direct effect of ACEs on M/ADs (β = 0.36, SE = 0.03, *p* <0.001), with individuals who experienced more ACEs being more likely to develop a M/AD. There was also a significant, albeit small, indirect effect operating through SDs (β = 0.04, SE = 0.01, *p* <0.001), which accounted for 10% (0.041/0.397) of the total effect of ACEs on M/ADs.

### Genetic Moderation and Moderated Mediation

<u>African Ancestry (AA).</u> Among AA individuals, the direct effect of ACEs on SD was moderated by the SUD PRS (β = -0.15, *p*<0.001), with a smaller effect of ACEs on SD for those with a higher SUD PRS (see Table 5 and Figure 2, Panel A). There was no evidence for moderated mediation in the forward or reverse models among AA individuals (see Table 5 and Figure 2).

**Table 5.** Results of moderated mediation models.

|  | African Ancestry |  |  | European Ancestry |  |  |
| --- | --- | --- | --- | --- | --- | --- |
| | $\beta$ | SE | p-value | $\beta$ | SE | p-value |
| ***Forward Model: Adverse Childhood Events (ACEs)→Mood and Anxiety Disorders (M/ADs)→Substance Dependence (SD)<sup>1</sup>*** |  |  |  |  |  |  |
| ACEs →M/ADs | 0.797 | 0.009 | <0.001 | 0.791 | 0.009 | <0.001 |
| ACEs x M/AD PRS | 0.016 | 0.012 | 0.181 | 0.018 | 0.011 | 0.106 |
| M/ADs→SDs | 0.204 | 0.024 | <0.001 | 0.170 | 0.019 | <0.001 |
| ACEs→SDs | 0.266 | 0.024 | <0.001 | 0.350 | 0.020 | <0.001 |
| ACEs x SUD PRS | -0.145 | 0.014 | <0.001 | -0.209 | 0.012 | <0.001 |
| M/ADs x SUD PRS | -0.007 | 0.014 | 0.596 | -0.009 | 0.011 | 0.405 |
| ***Reverse Model: Adverse Childhood Events (ACEs)→Substance Dependence (SD)→Mood and Anxiety Disorders (M/ADs)<sup>2</sup>*** |  |  |  |  |  |  |
| ACEs→SDs | 0.509 | 0.024 | <0.001 | 0.328 | 0.014 | <0.001 |
| ACEs x SUD PRS | 0.013 | 0.023 | 0.552 | -0.056 | 0.016 | <0.001 |
| SDs→M/ADs | 0.086 | 0.010 | <0.001 | 0.184 | 0.020 | <0.001 |
| ACEs→M/ADs | 0.530 | 0.012 | <0.001 | 0.674 | 0.013 | <0.001 |
| ACEs x M/AD PRS | 0.005 | 0.013 | 0.676 | -0.009 | 0.018 | 0.616 |
| SDs x M/AD PRS | 0.010 | 0.008 | 0.236 | -0.001 | 0.016 | 0.970 |

*Note:* Models included sex, age, marital status, years of education, employment status, and income as covariates. SE = standard error, CI = confidence interval, M/AD PRS = mood and anxiety disorder polygenic risk score, SUD PRS = substance use disorder polygenic risk score.
<sup>1</sup>Model was restricted to participants with an age of onset for first substance dependence diagnosis at or after their age of onset for a first mood/anxiety disorder diagnosis. African ancestry, n=1,200; European ancestry, n=1,809.
<sup>2</sup>Model was restricted to participants with an age of onset of a first mood/anxiety disorder at or later than the age of onset for a first substance dependence diagnosis. African ancestry, n=1,164; European ancestry, n=1,539.

<u>European Ancestry (EA).</u> Among EA individuals, consistent with results in AA individuals, the direct effect of ACEs on SD was moderated by the SUD PRS (β= -0.21, *p*<0.001), where again there was a smaller effect of ACEs on SD for those with a higher SUD PRS. There was no significant moderated mediation in this model (see Table 5 and Figure 2, Panel A). However, in the reverse or substance-induced model, there was evidence for moderated mediation of path a (from ACEs to SD; β= -0.06, *p*<0.001), where again higher genetic risk for SUD was associated with a smaller effect of ACEs on SD risk. There was no evidence that genetic risk moderated any other paths (see Table 5 and Figure 2, Panel B).

## DISCUSSION

Consistent with our hypotheses, there were both direct and indirect effects of ACEs on SDs and M/ADs. In a sample more than double the size of our previous study sample,^14^ we replicated our earlier findings that the effect of ACEs on SD risk was mediated by M/ADs. Leveraging the power of the larger sample, we extended these findings by examining effects in the reverse model—i.e., in which the effects of ACEs on M/ADs were mediated by SD risk. Although we found both direct and indirect effects in both the forward and reverse models—underscoring the bidirectional nature of the relationship between M/AD and SD—the indirect effect in the forward model was greater than that in the reverse model, providing greater support for the self-medication hypothesis than for substance-induced mood/anxiety disorders.

The self-medication hypothesis^15–17^, first posited in 1997, states that psychopathology is accompanied by aversive internal states that motivate individuals to use psychoactive substances to relieve them^57^. Research has shown that ACEs negatively impact emotion regulation skills^58,59^, which help individuals modulate distressing emotions in adaptive ways. These individuals may develop SDs following repeated attempts to alleviate troubling mood states and distressing emotions that result from exposure to ACEs by using substances. In contrast, the development of M/ADs following ACEs was largely due to direct effects, with mediation through SDs accounting for only 10% of the overall effect. This indirect pathway represents substance-induced M/ADs, which have been posited by some to account for a substantial amount of the comorbidity between SUDs and psychiatric disorders^60^. In this theory, M/ADs in the context of SUDs are temporary conditions that occur largely due to intoxication or withdrawal^60^. In contrast to M/ADs that develop from self-medication, those that are substance-induced resolve with abstinence from substance use^61^. Although our results reflect bidirectional pathways to M/ADs among individuals who experience ACEs, we found less support for the substance-induced pathway than the self-medication pathway. Thus, M/ADs among individuals who experienced ACEs are more likely to be a direct product of these early events.

These findings highlight potential opportunities to improve treatment outcomes among individuals who have experienced ACEs. Results of both our forward and reverse models highlight the importance of preventing or reducing young people’s exposure to ACEs as a means of decreasing the risk of both M/ADs and SDs. Improved screening efforts within healthcare and school settings^62,63^ and the facilitation of community cohesion^64,65^ show promise in mitigating the substantial public health burden of ACEs. Although substance misuse is often seen as a more pressing problem clinically, targeting premorbid M/ADs among individuals who use substances may help to reduce the intensity of use and thereby prevent the development of an SUD, even if the underlying M/AD does not remit fully. Whereas individuals in treatment for an SUD report rates of exposure to ACEs as high as 100%^66–68^, providing clinical interventions that target adaptive mood regulation to this patient population could help to reduce the impact of ACEs on psychosocial functioning^69^, improve retention in SUD treatment programs^70^, and reduce rates of relapse^71^.

Although as hypothesized, genetic risk for SUD moderated the effects of ACEs on the development of an SD, i.e., a GxE effect, it was in a direction opposite our hypothesis. Rather than an interaction effect consistent with the diathesis-stress model^72^, we found the association between ACEs and SDs to be *lower* among individuals with a higher SUD PRS. This suggests, first, that a higher genetic liability for SUD has a greater impact on the likelihood of an SD than an environmental (ACEs) factor. Thus, exposure to ACEs contributes more to SUD risk in individuals who are not highly genetically predisposed to develop an SUD. Taken together, this provides evidence of two different mechanisms by which an individual may develop an SUD following exposure to ACEs, which depend on the magnitude of the individual’s genetic liability to SUDs.

### Limitations and Strengths

This study has limitations. First, we chose to include PTSD in our latent M/AD factor because it is classified as an anxiety disorder in DSM-IV, the diagnostic system on which all psychiatric diagnoses in the Yale-Penn sample are based. This could have inflated the association between ACEs and M/ADs, because several of the ACEs (e.g., physical abuse or sexual abuse) are index traumas for PTSD. Second, there was greater heterogeneity in the M/AD latent variable than the SD latent variable, as evidenced by the lower trait loadings. This, combined with the smaller discovery GWAS for M/ADs ^36–44^ than for SUDs, particularly among African-ancestry individuals,^45–47,49^ may have contributed to the failure to detect moderated mediation effects for the M/AD PRS. Among African-ancestry individuals, the smaller discovery samples also limited statistical power of the SD latent genetic factor and PRS. Finally, although we used self-reported ages of onset to ensure the temporal precedence required for modeling mediation effects, the data were reported retrospectively, which could have introduced bias in the selection of individuals for the forward and reverse models. The sensitivity analysis in which we removed individuals who experienced their first M/AD and SD in the same year showed no substantive effect on model outcomes. This suggests that the findings are generally robust to errors in recall of age of onset of the disorders.

Despite these limitations, the study has key strengths, including its large discovery and target sample sizes, ability to examine mediation in two models of theoretical importance, and findings of genetic moderation in both AA and EA individuals. The use of gSEM to leverage the considerable overlap in SUDs and M/ADs to construct ancestry-specific latent genetic factors and then subject them to GWAS likely yielded greater statistical power than a single GWAS could provide. Such multivariate approaches could facilitate more equitable application of findings from genetic studies by enhancing power to detect effects even when discovery GWAS are of modest size, which is often the case for samples of non-European ancestry.

### Conclusions

Although we found support for both the self-medication hypothesis of SD and substance-induced M/ADs, there was greater support for the former. In fact, 90% of the risk of M/ADs was explained by the direct effect of ACEs. Interestingly, rather than heightening the effect of ACEs on SDs, among individuals with higher SUD polygenic risk, ACEs had less of an effect on SD risk, suggesting that the effect of genetic liability exceeded the impact of the environmental risk factor. In contrast, among individuals at lower genetic risk for SUDs, ACEs appear more likely to precipitate the development of an SD. PRS for M/ADs, however, did not moderate the effect of ACEs on M/ADs, indicating that the risk of developing these disorders is elevated by exposure to these childhood events, regardless of genetic liability. These findings argue for universal screening for ACEs and trauma-informed interventions in schools and communities to reduce the substantial impact of ACEs on risk for both M/ADs and SDs.

## Data Availability

All data produced in the present study are available upon reasonable request to the authors

## Acknowledgment

This work was supported by the Veterans Integrated Service Network 4 Mental Illness Research, Education and Clinical Center and by grants AA028292 to RLK and IK2 CX002336 to EEH.

