## supplementary figures for "Adverse Childhood Events, Mood and Anxiety Disorders, and Substance Dependence: Gene X Environment Effects and Moderated Mediation"

**Supplementary Figure 1. Factor loadings for the adverse childhood events (ACEs) latent variable.**

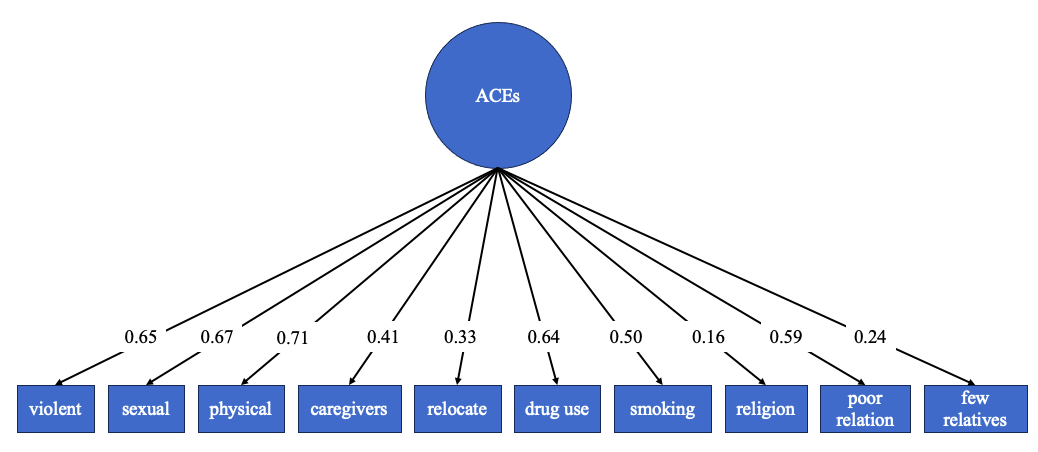

**Supplementary Figure 2. Factor loadings for the substance dependence latent variable.**

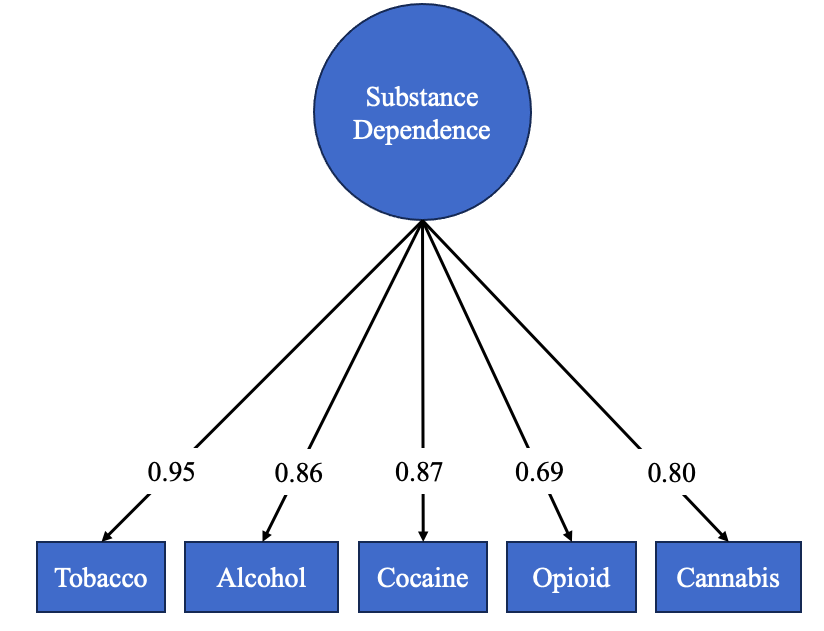

**Supplementary Figure 3. Factor loadings for the mood and anxiety disorders latent variable.**

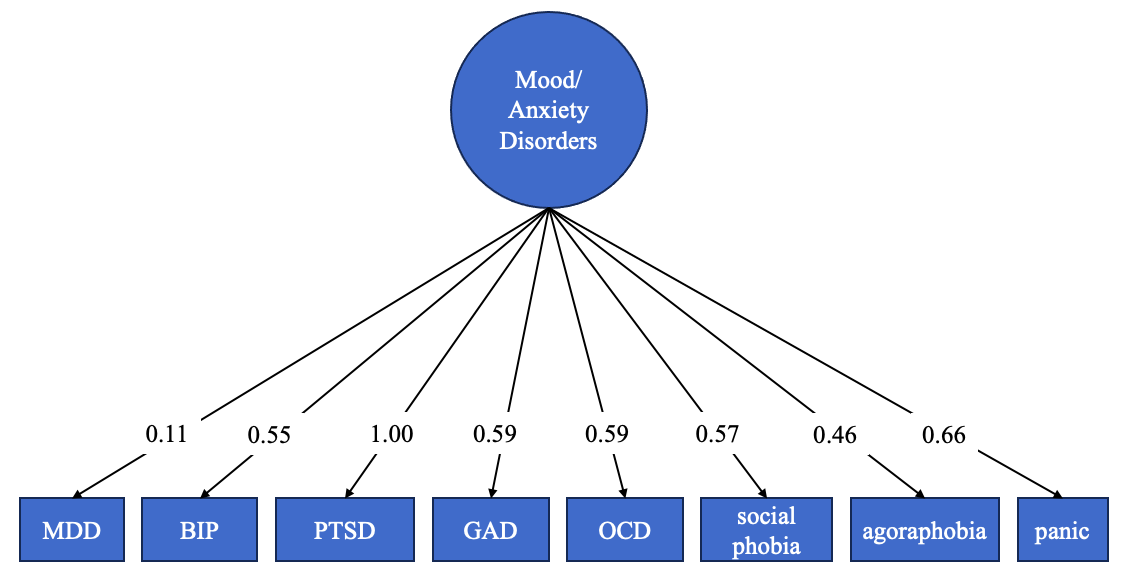

**Supplementary Figure 4. Mood and anxiety trait loadings onto common genetic factor, AFR ancestry**

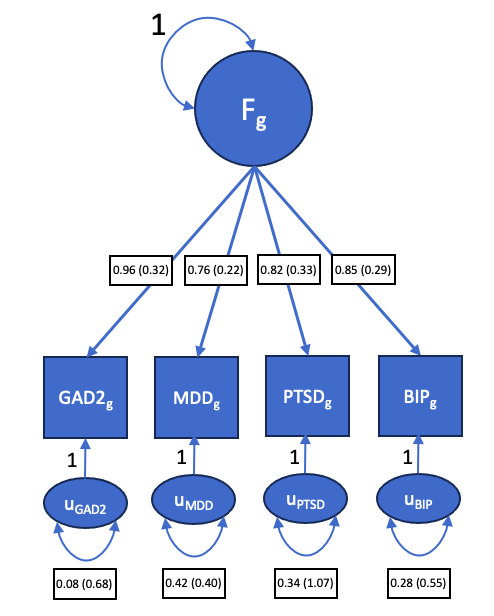

The common genetic factor was produced using GenomicSEM-0.0.5c from GWAS of each input trait shown in the diagram. Because African ancestry reference files are not provided with the GenomicSEM-0.0.5c suite, we used LD scores from the Pan-UK Biobank (https://pan.ukbb.broadinstitute.org. 2020.) to compute LD matrices and correlations. Analysis was then performed after filtering SNPs to include those with MAF>0.01 in the 1000G African ancestry population (https://doi.org/10.1093/nar/gkz836). Values attached to arrows from F_g_ to trait represent loading of the trait onto the common factor. Values at the bottom represent residual variance in each trait that is unexplained by the common genetic factor. Standard error values are in parentheses. GAD2= generalized anxiety disorder; MDD=Major depressive disorder; PTSD=posttraumatic stress disorder; BIP=bipolar disorder

**Supplementary Figure 5. Mood and anxiety trait loadings onto common genetic factor, EUR ancestry**

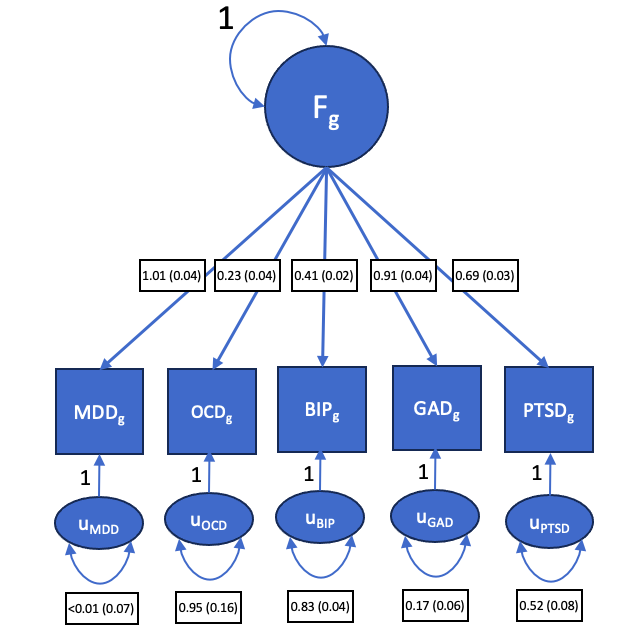

The common genetic factor was produced using GenomicSEM-0.0.5c from GWAS of each input trait shown in the diagram. The GAD trait was constructed from three GWAS of anxiety-related traits that were jointly analyzed using MTAG (methods). Values attached to arrows from F_g_ to trait represent loading of trait onto the common factor. Bottom values represent residual variance in each trait that is unexplained by the common genetic factor. Standard error values are in parentheses. GAD2=generalized anxiety disorder; MDD=Major depressive disorder; PTSD=posttraumatic stress disorder; BIP=bipolar disorder

**Supplementary Figure 6. Substance use disorder trait loadings onto common genetic factor, AFR ancestry**

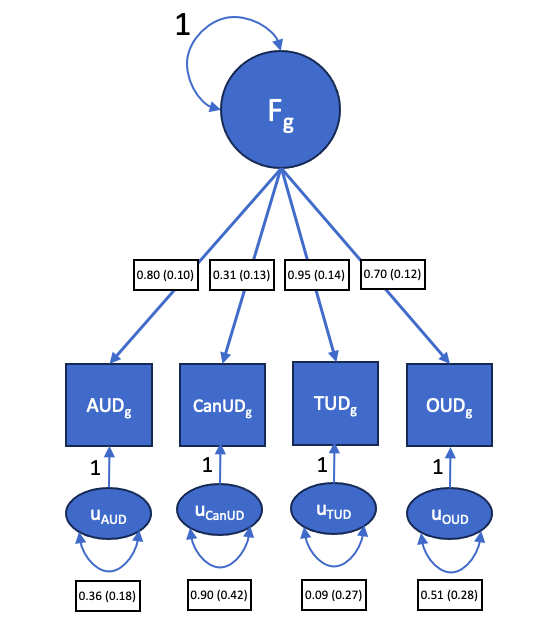

The common genetic factor was produced using GenomicSEM-0.0.5c from GWAS of each input trait shown in the diagram. Because African ancestry reference files are not provided with the GenomicSEM-0.0.5c suite, we used LD scores from the Pan-UK Biobank (https://pan.ukbb.broadinstitute.org. 2020.) to compute LD matrices and correlations. Analysis was then performed after filtering SNPs to include those with MAF>0.01 in the 1000G African ancestry population (https://doi.org/10.1093/nar/gkz836). Values attached to arrows from F_g_ to trait represent loading of trait onto the common factor. Bottom values represent residual variance in each trait that is unexplained by the common genetic factor. Standard error values are in parentheses.

**Supplementary Figure 7. Substance use disorder trait loadings onto common genetic factor, EUR ancestry**

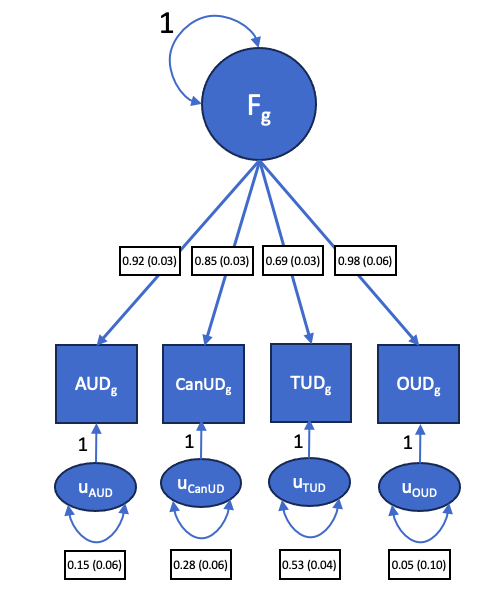

The common genetic factor was produced using GenomicSEM-0.0.5c from GWAS of each input trait shown in the diagram. Values attached to arrows from F_g_ to trait represent loading of trait onto the common factor. Bottom values represent residual variance in each trait that is unexplained by the common genetic factor. Standard error values are in parentheses.
